# Perception of game-based rehabilitation in upper-limb prosthetic training: a survey of users and researchers

**DOI:** 10.1101/2020.09.03.20186718

**Authors:** Christian Garske, Matthew Dyson, Sigrid Dupan, Kianoush Nazarpour

## Abstract

**Background:** Serious games have been investigated for their use in multiple forms of rehabilitation for decades. The rising trend to use games for physical fitness in more recent years has also provided more options and garnered more interest for their use in physical rehabilitation and motor learning. In this paper, we report the results of an opinion survey of serious games in upper limb prosthetic training.

**Objective:** This study investigates and contrasts the expectations and preferences for game based prosthetic rehabilitation of people with limb difference and researchers.

**Methods:** Both participant groups answered open and closed questions as well as a questionnaire to assess their user types. The distribution of the user types was compared with a Pearson’s *χ*^2^ test against a sample population. The data was analysed with the thematic framework method; answers fell within the themes of usability, training, and game design. Researchers shared their views on current challenges and what could be done to tackle these.

**Results:** A total of 14 people with limb difference and 12 researchers took part in this survey. The open questions resulted in an overview of the different views on prosthetic training games between the groups. The user types of people with limb difference and researchers were both significantly different from the sample population with *χ*^2^=12.31 and *χ*^2^=26.50, respectively.

**Conclusions:** We found that the respondents show a general willingness and tentative optimism towards the topic, but also acknowledge hurdles limiting the adoption of these games by both clinics and users. Results indicate a noteworthy difference between researchers and limb different people in their game preferences, which could lead to design choices that do not represent the target audience. Furthermore, focus on long-term in-home experiments is expected to shed more light onto the validity of games in upper limb prosthetic rehabilitation.

## Introduction

Serious games have been shown to enhance the outcome of movement rehabilitation after stroke or cerebral palsy [1,2]. Computer games have also been researched for upper-limb prosthetic control training since the early 1990s [3–14]. While the field has received considerably more attention over recent years, it has yet to succeed in finding general proof for successful transfer of training myoelectric control in the games to an increase in prosthetic control ability [15]. For instance, transfer has only been found in training a strongly ADL-relevant task in a virtual environment [15].

The skills required to play a game can be learned with practice. A serious game incorporating training exercises consistent with a rehabilitation regimen allows the numerous repetitions of exercises necessary during rehabilitation therapy to take place. Nonetheless, it is hypothesised that the training needs to be engaging to ensure regular and consistent adherence to the exercises [12,16]. As such, current prosthetic training games revolve around two core themes: the engagement of the player [7,8,12,14,16–20] and the skill transfer from the game to prosthetic use [15,20,21]. These two objectives are pursued with two types of *myo-games*: 1) games that are based on existing game platforms with input adjusted to the electromyogram (EMG) signals [4,6] and 2) completely novel games around specific training goals [9,20].

Few researchers specifically acknowledge the diversity of the target audience and its effect on the success of myo-games [16,17,22,23]. Even fewer have attempted to address this difference in preferences in their developed games and take the views of people with limb difference into account before starting the development process. A notable exception is the work of Tabor *et al*. [16] who collected qualitative feedback from a small testing group and incorporated the suggestions into their game design. Because of the diversity in the population, where people have varying definitions of fun aspects and expectations for games, multiple attempts have been made to categorise people into user types [24–26]. Professional game development is based on the psychology behind those types of gamers and the choice of the appropriate game design elements fitting for the target audience. However, academic game development is typically carried out over a relatively short period of time and by a small, non-specialist team, which is in stark contrast to the years of development time often carried out by a large and highly specialised team that go into modern games. This means that the decision-making in these games is potentially subject to the preferences of a small team that does not generally reflect the preferences of the target audience.

We hypothesised that there are considerable differences between the views of prosthesis end users and the researchers with regards to engaging aspects of a game. To test this hypothesis we created a survey and sought to determine the focus points of each of these groups. In addition, we included a user type questionnaire to ascertain the distribution of each participant group for comparison. Utilising such a user type distribution can furthermore deliver useful information to lead general design choices in the game development. It could also be used for presets that emphasize certain game design elements over others for increased user engagement. Furthermore, this survey aimed to identify other challenges than a potential disparity in game preferences that the community of researchers could have to face on the path of game-base upper limb prosthetic rehabilitation. This work adds the opinions of researchers and limb different people about games in upper limb rehabilitation to the research that has been conducted on the opinions of clinicians [27].

## Methods

The study was approved by the University Ethics Committee of Newcastle University under the reference number 905/2020. The survey ran from February 2020 until May 2020. Participants were either people with upper-limb difference or researchers who are active in the research of games for prosthetic training or in prosthetic research in general. All participants gave their consent by filling out and submitting the survey as stipulated on the first page of the survey form.

The recruitment of this study was conducted predominately online via personal contacts. Additional outreach was done via social media and by contacting charities in the UK that are involved with people with limb difference. Specifically, the survey was sent out to main and local branches of 13 different charities as well as 40 researchers involved in upper limb prosthetic research. The inclusion criterion for people with limb difference was the absence of the upper-limb, irrespective of level, side (uni- or bilateral) or whether they use a prosthesis or not. Fourteen people with limb difference and twelve researchers filled out the survey. An overview of the demographic data of the participants can be seen in table 1 and table 2. We chose an online survey and expected that the online nature would increase the number of people willing to participate due to ease of access. Additionally, the online survey offered the participants time to think about their answers without the pressure of coming up with an answer on the spot. The survey was developed in English and was not altered over the course of the study. The participants were given the option to contact the authors in case they did not want or were not able to fill out the survey online. No participant made use of this option.

**Table 1.** Table shows participants’ gender (self-identified, M: Male, F: Female); age group; type of limb difference (A: Amputation, C: Congenital); side and level of limb difference, prosthesis use, and former participation in research.

| Gender | Age Group | Type | Side | Level | Use | Participation |
| --- | --- | --- | --- | --- | --- | --- |
| F | 31-40 | A | Dominant | Above Elbow | No, but interested | Yes |
| M | 31-40 | C | Dominant | Below Elbow | Former | No |
| F | 41-50 | A | Both | Below Elbow | Tried | No |
| F | 41-50 | A | Non-Dominant | At Shoulder | No, but interested | No |
| M | 41-50 | C | Non-Dominant | Below Elbow | Active | Yes |
| F | 41-50 | A | Dominant | Below Elbow | No, but interested | No |
| M | 41-50 | C | Non-Dominant | Below Elbow | Active | No |
| F | 41-50 | A | Dominant | Above Elbow | Tried | Yes |
| M | 51-60 | A | Dominant | Below Elbow | Active | Yes |
| F | 51-60 | A | Dominant | Below Elbow | No, but interested | Yes |
| F | 51-60 | C | Non-Dominant | At Wrist | No, no interest | Yes |
| M | 51-60 | C | Non-Dominant | Below Elbow | Active | Yes |
| M | 51-60 | A | Dominant | Below Elbow | No, but interested | No |
| M | 61-70 | A | Non-Dominant | Below Elbow | Active | No |

**Table 2.**
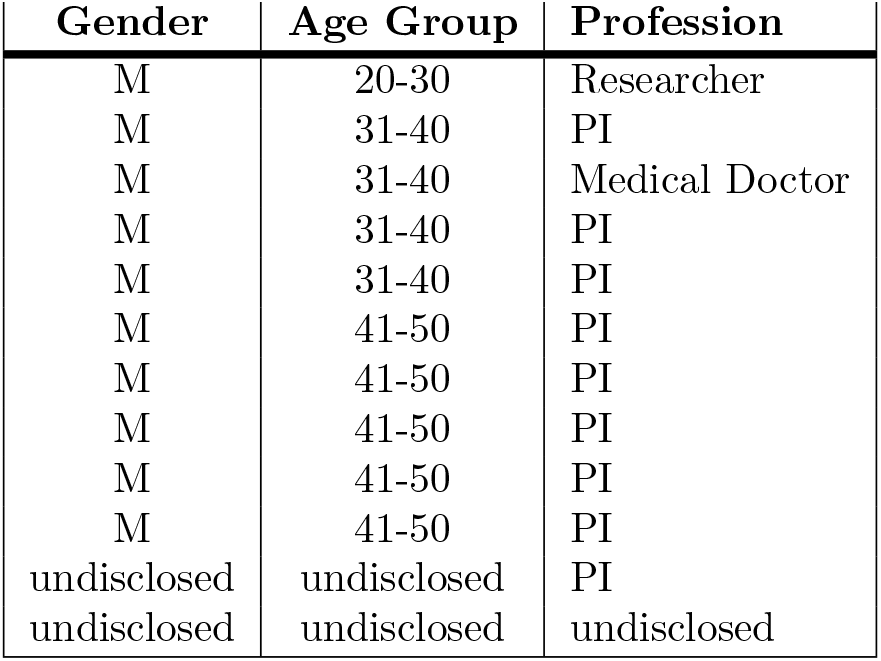
Table shows researchers’ gender (self-identified, M: Male, F: Female); age group; and academic level (PI: Principal Investigator).

The survey first introduced the general aim of the study and the contact information of the first and last author and the Data Protection Officer of Newcastle University. The survey asked for general demographics and, in case of people with limb difference, for an anamnesis with regards to their limb. This was followed by a user type questionnaire, originally developed by Tondello *et al*. [26]. The survey concluded with open questions about the participants’ preferences and opinions with regards to games in general and games in prosthetic training specifically. The researchers were asked to answer additional questions concerning challenges in this field of research.

## Data analysis

The results of the user type questionnaire were processed with MATLAB. A goodness of fit test using Pearson’s chi-squared test with a significance level of *α* = 0.05 was conducted for both participant groups compared to the distribution published by Tondello *et al*. [26]. This test was chosen to identify potential differences between the distribution of user types of the participant groups and the distribution of a larger sample population.

For the analysis of the resulting data for the open questions of the survey, we applied the Thematic Framework approach [28]. This approach consists of five steps:

1. Familiarisation: All authors familiarised themselves with the collected data. The first author created an initial theme set, which was discussed and agreed upon by all authors.
2. Identifying a thematic framework: The first author created a set of subthemes for the data set. This was approved by the last author for use in the next steps. The full set of themes can be found in table 3.
3. Indexing: All authors coded the interview data independently. These were discussed between all authors until a consensus was reached.
4. Charting: The data was sorted by themes and subthemes by the first author.
5. Mapping and interpretation: The first author summarised and interpreted the charted data according to the themes.

**Table 3.** Thematic Framework. Table shows the proportion of responses per group in % and rounded to the first decimal place, followed by total number in brackets. The percentages are further split into the respective subthemes.

| Themes | Limb Different | Researchers |
| --- | --- | --- |
| **Usability** | **15.4** (6) | **10.2** (11) |
| Accessibility | 10.3 (4) | 3.7 (4) |
| Data Management | 0 (0) | 1.8 (2) |
| Hardware | 5.1 (2) | 4.6 (5) |
| **Training** | **35.9** (14) | **35.2** (38) |
| Muscle Development/Control | 10.3 (4) | 3.7 (4) |
| Prosthetic Ability | 17.9 (7) | 13.9 (15) |
| Additional Benefits (Users) | 5.1 (2) | 7.4 (8) |
| Clinical & Research Benefits | 2.6 (1) | 3.7 (4) |
| Education | 0 (0) | 3.7 (4) |
| Feedback | 0 (0) | 2.8 (3) |
| **Game** | **48.7** (19) | **29.6** (32) |
| Affect | 12.8 (5) | 15.7 (17) |
| Personalisation | 7.7 (3) | 6.5 (7) |
| Social Aspects | 0 (0) | 0.9 (1) |
| Mechanics | 28.2 (11) | 6.5 (7) |
| **Challenges** | **0** (0) | **25.0** (27) |
| Justification/Reasoning | - - | 10.2 (11) |
| Design/Development | - - | 9.3 (10) |
| Involvement | - - | 1.8 (2) |
| Recognition | - - | 3.7 (4) |
| Total | 100 (39) | 100 (108) |

## Results

Fourteen people with limb difference and twelve researchers in the prosthetic field participated in the survey (see tables 1 and 2, respectively). Seven of the people with limb difference had experience with studies pertaining rehabilitation with a type of computer-based device prior to the survey.

An overview of the themes identified from the survey data is presented in table 3. In the following sections the responses of the survey participants are presented. The texts in the *italic* font are direct quotes from the participants. Results are reported thematically: usability, training, game design and challenges. The raw survey results are accessible in supporting material.

## Usability

The majority of the participants, both limb different people and researchers, indicated that they believed people in prosthetic training would use a game-based training at home, figure 1A. Three out of the 14 limb different people and five out of the 12 researchers cautioned that factors predominately concerning usability and game design can affect adoption, as usability is one of the issues as to why former gamers have stopped playing virtual games.

> *I used to play an Atari, as it had joystick controls, but I struggle with handsets, so [I have] not played for some time*.

**Figure 1.**
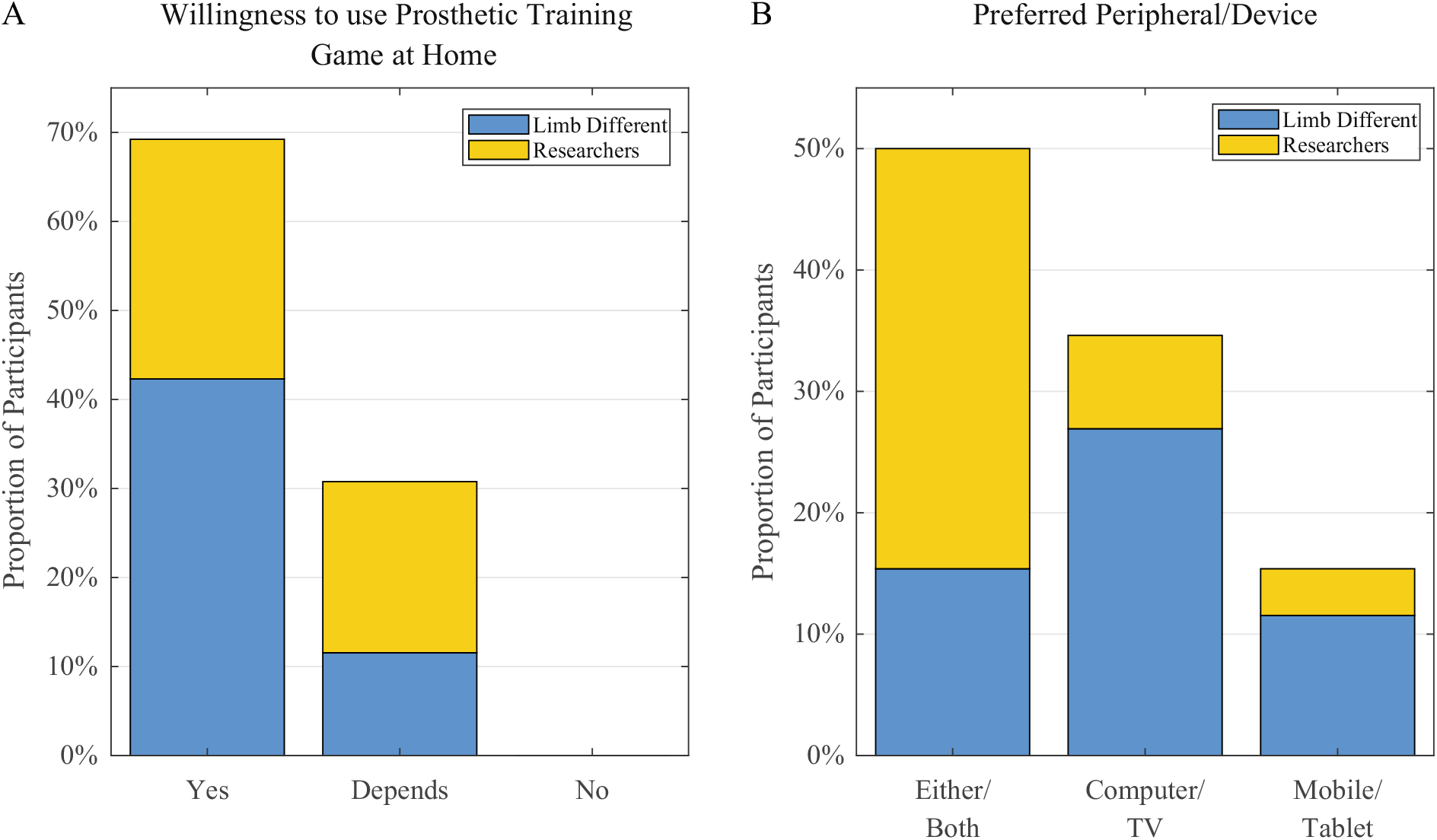
Participant response proportions for (A) their willingness to use a game-based prosthetic training tool at their own home and (B) their preferred peripheral or device for such a game.

In addition, participants pointed out that such a tool should prove easy and robust to use both in set-up as well as in the actual use of the tool.

An additional concern was raised with regards to the compatibility of the software with potentially existing hardware at home. One participant said:

> *[Requirements for game-based prosthetic training include] providing a setup that is easy to use and robust; ensuring a game framework that allows people to use their own devices*.

Creating game framework that would work cross-platform with potentially outdated hardware is a significant challenge for the myoelectric control research community. A step in this direction could be the development of online gaming platforms as mentioned by one of the researchers. Furthermore, researchers indicated the importance of the exchange of code and knowledge within the community.

The preferred choice of platforms and peripherals, the input and output devices with which the game is played, was more distinct in the case of the participants with limb difference than the researchers. Figure 1B shows that the majority of limb different people would prefer a computer or TV screen. However, researchers showed no clear preference for the mobile over fixed screen options.

## Training

The topics connected to the main theme of training were grouped into muscle development and control; prosthetic ability; additional benefits for users, clinicians and research; education; and feedback.

The answers concerning muscle development and control mostly centred on the participants’ hopes and expectations. People with limb difference expect the game to help developing the musculature in their remaining limb and improved prosthetic dexterity with these muscles. For instance, a limb different participant wished for a game that highlighted:

> *my ability to use advanced prosthetics*.

Another participant with limb difference would like to see a game made that also helps to reduce the phenomenon of phantom limb pain.

Researchers additionally specified their hopes in the direction of accelerated learning speeds, the training of ADL-related tasks and the transfer of the skills learned in the game to prosthesis use. One of the researchers stated that any tool created should be based on the needs of the patients as well as clinicians. Outside of the physical benefits, researchers pointed out that a game-based tool could let the user share data with other users, if they wanted to, and also potentially lead to a reduced need for input through in-clinic appointments.

In addition, a training game could support therapists in assessing the patients’ ability. One researcher indicated:

> *In the end the games should also relieve the therapists from time spending on rehabilitation. Moreover, the games should be helpful as an assessment tool for the appropriateness of a prosthesis for a given patient*.

The need of personalised feedback for the users was raised by a couple of researchers. They envisioned the benefits of regular and individualised training progress feedback by which users as well as therapists can track their skill progression properly.

## Game Design

The answers of the participants directly relating to design choices for the game were grouped under the main theme of game design. The subtheme of affect included all answers that aimed at an emotional response of the user; personalisation contains in-game control of design for the player; social aspects revolve around the possibility of interaction with other players; and answers in the mechanics subtheme pertain to ideas for the mechanical features of the game.

With regards to the affective aspect of the design, participants of both groups indicated that the game has to be engaging for in-home adaption. For instance, one researcher points out that rehabilitation games should not only be interesting in the short term, but also be able to secure the long-term engagement of the user. However, engaging the user to a sufficient level was recognised as a challenge.

Furthermore researchers say the game should be attractive to users, though it could be hard to appeal to the wide range of potential users. A range of games or in-game options could be beneficial due to the differing motivating factors. A degree of immersion was wished for by a limb different participant and another pointed out the potential of an *empowering portrayal of life post-amputation* in the game, e.g.:

> *That the game would be so immersive that the participant would not realise they are training their remnant muscles for optimal EMG based control*.

Interestingly, one limb different participant suggested an *End of world* setting. Because this might not feel appealing for all users, the setting should be customisable by the player. Some participants from the limb different group wanted to see a relatable character using a prosthesis in the game, tying in with the empowering effect mentioned earlier. This affords a viable option for personalisation.

> *Perhaps the protagonist could be a prosthetic wearer […]*.

The variety of preferences in the thematic setting of the game can be seen in figure 2A. These are results of a multiple choice question and therefore do not add up to 100%.

**Figure 2.**
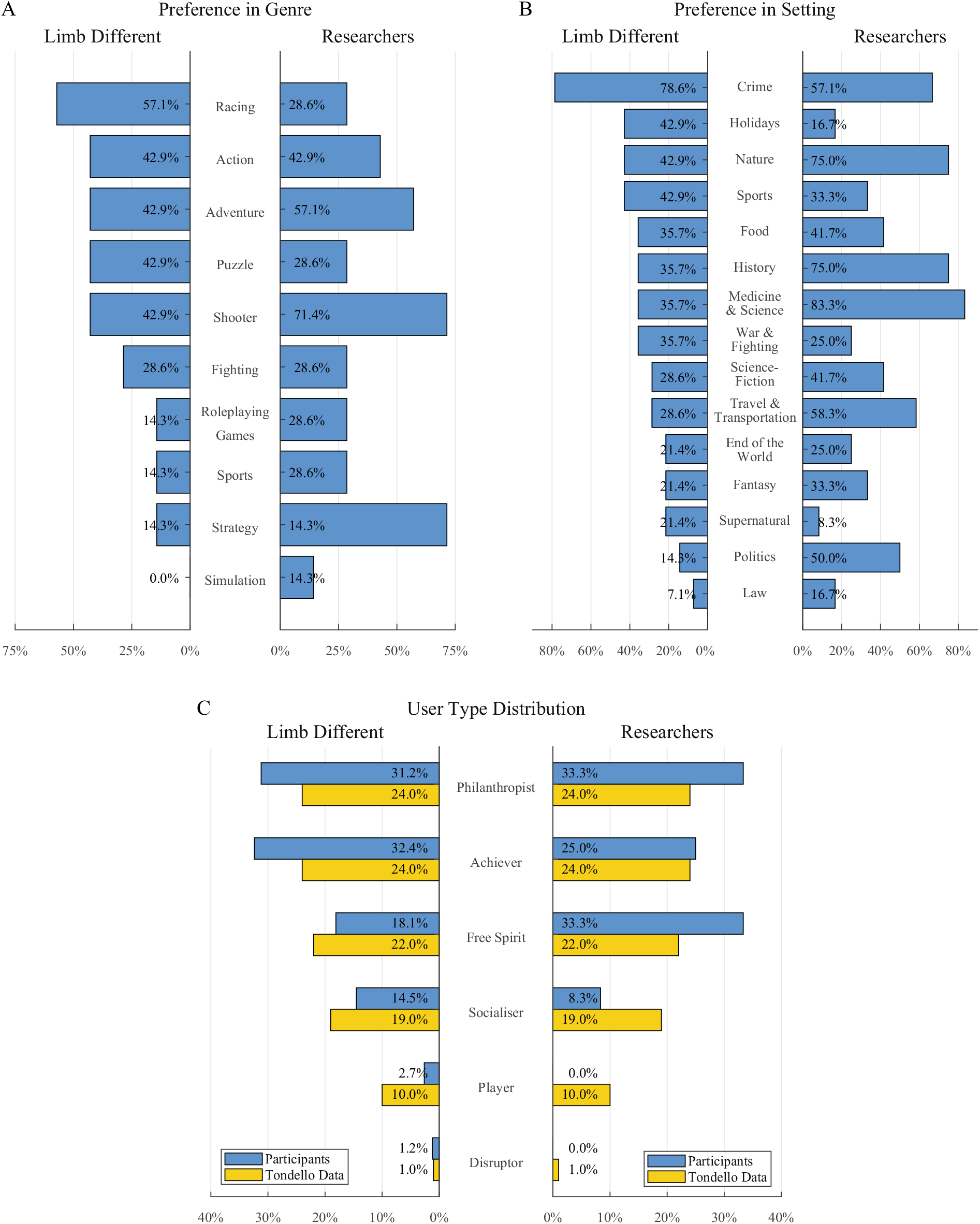
Results of the multiple choice questions regarding (A) the participants’ preference in genre and (B) preference in setting. These do not add up to 100%. Furthermore, (C) the results of the evaluation of the user type questionnaire of the participants (blue) contrasted with the sample population in Tondello *et al*. [26].

The mechanics of a game-based prosthetic training tool is predominately addressed with respect to the game genre. Participants of the limb different group identified a variety of different genres as desirable, including quiz and puzzle games, but also adventure, shooter and fighting games as well as horror games. One participant mentioned more specific activities like camping, fishing and shooting. A researcher argued against the use of war and fighting mechanics in clinical settings. The different genre preferences of the participants can be found in figure 2B. As before, these results do not add up to 100%.

An additional influence on the type of game and the game mechanics involved can be the type of gameplay the user prefers. As it cannot be assumed that a user is an active or former gamer and therefore knows what they look for in a game, an assessment of the user types using the Hexad Scale [26] was conducted in this survey. The outcome of this assessment both for limb different people and researchers can be seen in figure 2C. Among the limb different people, the philanthropist and the achiever user type are tied as clearly the most common types, whereas the player and the disruptor are the least commonly occurring. None of the researchers were grouped into the players and disruptors, which is similar to them being the least common type in the other group. We observed a notable difference in the overall distribution of user types to the limb different people. A Pearson’s *χ*^2^ test was conducted to test the statistical significance of this difference. Results showed that both groups are significantly different to the control group in Tondello *et al*. [26] at a significance level of *α* = 0.05. However, the result for the limb different people at *χ*^2^_*LD*_ = 12.31 are noticeably closer to the critical value of *χ*^2^ = 11.07 than the result of the researcher participant group of *χ*^2^_*LD*_ = 26.50. The group of researchers is even more different from that sample population (*α* = 0.005). This indicates that the group of limb different participants show a higher similarity to the sample population presented in Tondello *et al*. [26] than of the group of researchers do.

Apart from the genre, people with limb difference indicated that they would like a progressive and appropriate increase in difficulty, use both hands to play the game and for the game to motivate them to make enough repetitions of the trained arm to form habits. A researcher pointed out that the abstraction of the signals to rewarding or menacing game elements could be beneficial.

## Challenges

Only researchers participated in this part of the survey. They were asked to formulate their opinions on the challenges that the field of games in upper limb prosthetic rehabilitation faces. Additionally, they were invited to propose potential actions that could be undertaken by the community to address these challenges.

Part of the challenges mentioned by the researchers was the justification for the use or the development of serious games in prosthetic rehabilitation. The meaningful impact in terms of skill transfer to prosthetic use by myo-games must be investigated. This was stated not only for short term effects but also for long-term benefits when compared to other rehabilitation methods. The recognition of the difference between in-game improvement and actual benefit for prosthetic use was deemed not widely acknowledged yet.

> *However, most game studies focus only on in-game improvement. Now in-game improvement is a requirement for transfer to daily life performance. However, in-game improvement is not a sufficient requirement for transfer*.

Therefore researchers called for longitudinal and large-cohort studies in the field to show the appropriateness of the medium used and the transfer capabilities of - potentially only certain types of -games.

The development of these myo-games faces its own problems and challenges. They should make the benefits clearly visible for the user but at the same time make the training imperceptible by shifting the focus of the user away from the underlying reason for the training onto the task-specific in-game goals. Researchers indicated that the formation of bad habits to win the game by potentially compromising the training efficiency should be avoided. Therefore it was recommended to involve game developers in the process and to parallelise game development and transfer testing procedures; to not lose sight of either aspect. One researcher proposed the development of a knowledge and information sharing platform for myo-games. He also indicated that such a platform could lead to a wider and easier access of developed games for users at home with their existing hardware

Finally, the recognition of the value of myo-games is another key obstacle to tackle. Some people and especially certain age groups might dismiss games as frivolous and a waste of time. Both clinicians and patients might need to be convinced that a serious game for prosthetic rehabilitation could benefit them as well as it could potentially benefit research. For example, a researcher suggested:

> *Educating participants about serious games and why the time they spent playing is well spent […]*

Reviewing the number of mentions of the main themes, a clear separation of themes becomes visible. In figure 3 it can be seen that the main expectation of the participants for the prosthetic training lie within the training benefits that it should provide. The fewer mentions of game design topics indicate that the design is of concern but that the training aspect takes priority and should be the base minimum of any game-based training. The themes of training and game design almost have the same amount of mentions overall in this survey with 52 and 51 responses, respectively. In contrast to the clear focus on training in the expectations, the game design related themes were more split between preferences expectations. This suggests that many game design traits are considered desirable but not a necessary component, which is reflected in some research in the field of game-based prosthetic training.

**Figure 3.**
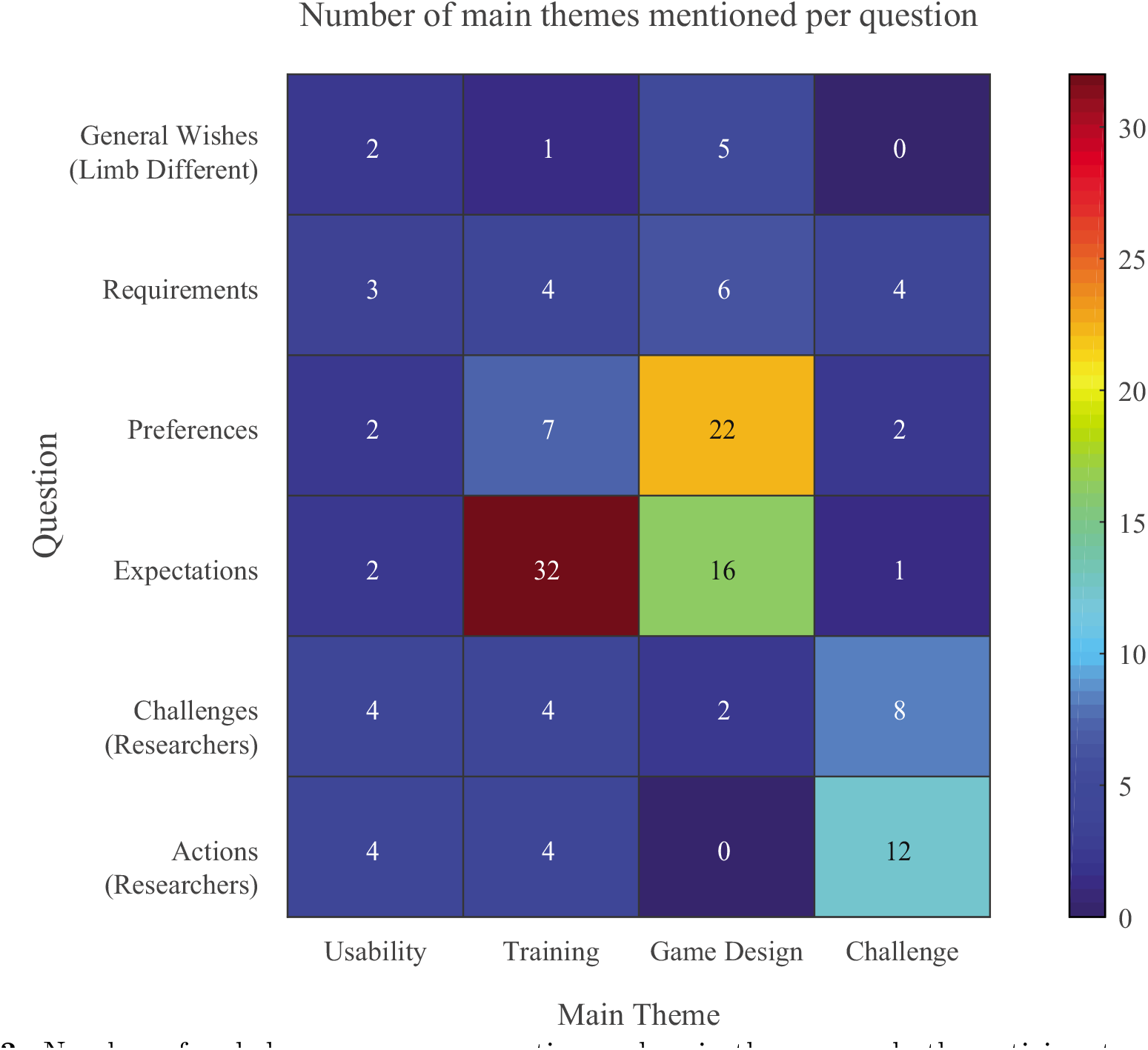
Number of coded responses per question and main theme over both participant groups.

## Discussion

The aim of this study was to determine the preferences, the expectations, and the views of both researchers and people with limb difference on game-based prosthetic training. The outcome of this survey indicates a general willingness and tentative optimism towards the topic. But for the wide adoption of myo-games, several scientific and engineering challenges should be addressed.

## Myo-games: opportunities and challenges

In the following, we will discuss all identified challenges in the context of the usability, training and game-design themes.

## Usability

With respect to hardware usability, the discontinuation of the commonly used Thalmic Myo Gesture Control armband [9,13,20], means there is now a lack of low-cost, easy to use EMG sensors for home use. Of the commercially-available dry EMG options suitable for game-based training, almost all require sensors to be accurately positioned using adhesives. As such, researchers are increasingly developing custom-built EMG acquisition solutions for game-based systems [14,18,29]. This approach does not scale well and is likely to contribute to slow translation of laboratory research to translational research with larger number of participants.

Accessibility of in-home myo-game software is also a key factor for overall usability. Accessibility must be balanced against client privacy and patient confidentiality, key points which therapists have identified as important in game based upper limb rehabilitation [27]. The small target population of upper-limb amputees and the niche nature of game-based rehabilitation mean cross-platform software with widespread hardware compatibility is not likely to be financially viable. The low cost of Android mobile devices, and the fact that they can be locked down to a restricted set of sandboxed applications, make this platform the most likely candidate for in-home use.

## Training

The topic of training with regards to goals was fairly unanimous between the limb different participants and the researchers. In current research, it appears to have mostly been assumed that an improvement in game control would translate readily to an improvement in prosthetic control as only in-game improvement or abstract control has been measured by most studies [4,6,8,12–14,20,30,31]. The question whether this transfer can happen and with which type of game this might happen has yet to be answered, as current research challenges the idea that a general myocontrol skill exists [10,32]. Although not only direct transfer could prove beneficial but also indirect improvement to learning the use of prosthesis, for example with respect to training speed, should be investigated. Therapists especially have expressed that gaming in therapy should be balance with other forms of therapy [27] and as such it might prove beneficial to examine then in conjunction.

The only testing of the effect on actual prosthetic skill in game-based prosthetic training research was conducted on the direct effect that the developed game would have on prosthetic skill [10,15]. We could not find any study that has conducted research if the training with a game prior to or alongside the actual training with a prosthesis could be beneficial to the learning process as opposed to an immediate effect on prosthetic skill. Prior training could be sensible as post-amputation, the site of surgery may not yet be ready for the fitting of a prosthetic socket. The British Society for Rehabilitation Medicine state that the fitting may be deferred to four to six weeks after the amputation [33]. In both the amputees and congenitally limb different people the muscle sites could need development before a prosthesis can be considered. Training alongside real life prosthetic training could add to the fine control without being reliant on the other arm musculature when those muscles are tired from the weight of the prosthesis.

Responses from the limb different participant group indicate that they would like a prosthetic training tool which makes rehabilitation feel less like rehabilitation. This was reflected in the call of the researchers for immersion in the game which can let the focus of the user shift from their limb and muscles to the task at hand, allowing the motions to become intuitive. This can also be found as part of the OPTIMAL (Optimizing Performance Through Intrinsic Motivation and Attention for Learning) theory of Wulf and Lewthwaite [34] stating that an external focus has beneficial effect on motor learning as well as the sense of accomplishment. According to that theory, the external focus as well as intrinsic motivation feed into a virtuous cycle of enhanced motor learning. However, the question how to incorporate these aspects most efficiently is yet to be answered.

## Game Design

One of the main points mentioned for the game design was the facilitation of both short- and long-term engagement. The potential users of a prosthetic training tool seem willing to take the leap to use it, but the appeal of such a novelty can quickly wane. It is the task of good game design to support the user by providing motivating game play for the whole training period. This again feeds into the aforementioned virtuous cycle according to Wulf and Lewthwaite [34].

The involvement of game developers would be a sensible path for research to take. This can be especially motivated as game developers have more experience in catering towards a specific audience with their games and which game design elements work best to keep the users motivated to play the game. Academic teams are likely to have different views on fun themes and activities to the general population based on the difference of the user types in this study. The inclusion of the design preferences of the potential end-users should also be considered by the researchers developing the games. Nonetheless, this gives rise to the question of how to achieve the effective targeting of the game in a so varied target audience. Working towards increasing the engagement of a game prior to knowing whether transfer will happen for this game could lead to fruitless efforts. However, it is possible that the engagement is a contributing factor in the transfer process and therefore worthy of further exploration.

As the current state of myo-game development is very diverse, it was expected that the opinions on the matters to focus on and the potential resolutions are just as varied.

## The survey: strengths and limitations

It is the first time a survey like this has been conducted in this field with these participant groups. We did not include clinicians in this survey as a previous study covers the therapists’ views on the use of video games in general upper limb rehabilitation [27]. The format of the study allowed the easy spread of the survey among the field. This led to the recruitment of 12 researchers and 14 limb different participants. However, the sample size of the survey turned out smaller than would be desirable for a good cross-section of the limb-different population as well as researchers. In an attempt to acquire many participants, this survey has been spread in various manners, but it is possible that survey fatigue has stopped people from participating. It could indicate that other means of interacting with limb different people might be advisable for future research. If a similar study was to be repeated in this format, we recommend collaborating on it with several research groups. This would provide a access to a larger group of researchers as well as people with limb difference.

Additionally, regarding the data set of the researchers, the clear majority of participants identifying as male could influence the outcome of the answers. Female researchers were included in the list the survey was distributed amongst. However, due to the general gender disequilibrium in the field this list already contained a higher percentage of male researchers. This could have an influence on the opinions and preference distribution of the researchers, as differences in gaming preferences have been identified in both gamer and non-gamer populations [35].

## The future of myo-games

In this study, the results indicate a higher level of similarity of the limb different participant group to the sample population in Tondello *et al*. [26] than of the researcher participant group. While the low number of participants does not make for conclusive evidence in this matter, it is still worth considering the implications of this dissimilarity. The influence of personal preferences and assumptions made by the researchers could have significant impact on the engaging and motivating aspects of the game they are developing. This could be mitigated by bringing in professional support from the game development sector or increase the collaboration with game experts within academia, who have more practical knowledge in catering a game experience towards a diverse target audience. However, as the market for serious games for upper limb prosthetic rehabilitation is fairly small and therefore the potential profit margin is small, including professional game developers in this research could prove challenging.

Furthermore, the effect of myo-games has mostly been investigated in short trials of up to a week in experimental scenarios at a university or at the home of the participant. A much improved assessment of the effect of these games in both the short and the long term as well as whether a significant level of transfer occurs could be achieved by conducting longer term home trials with limb different people. This would also provide more information about the engaging aspects of the game in the long term, i.e. if the feelings of the participants change due to the waning novelty of the game and if it becomes a chore. Inversely, if the attrition over a long-term study would prove to be significantly lower than comparable experiments, this could be an indicator for a positive effect of games to exercise and training adherence.

The current state of isolation due to the 2020 coronavirus pandemic poses many challenges for academic research, some of which may persist into the long term. Thus a shift in experimental procedures to the home environment of the participants could be a beneficial route to follow. Many of the technical challenges surrounding home based experimentation, such as precision timing, low latency networking and data security have already been widely addressed in gaming. More widespread adoption of gaming technology to facilitate the shift of experimentation to participants’ homes may provide an alternative route to bridge the gap between academic research and viable prosthesis training solutions.

## Data Availability

The data is available in the supplementary file.

## Acknowledgements

This work has been supported by the Leverhulme Doctoral Scholarship Programme in Behaviour Informatics (DS-2017–015), and the Engineering and Physical Sciences Research Council (EPSRC) via grants EP/R004242/1 and EP/M025594/1.

## Abbreviations

EMG: Electromyography

## Supporting information

S1

Text. Raw Results. (PDF)

